# Efficient mucosal antibody response to SARS-CoV-2 vaccination is induced in previously infected individuals

**DOI:** 10.1101/2021.12.06.21267352

**Authors:** Kaori Sano, Disha Bhavsar, Gagandeep Singh, Daniel Floda, Komal Srivastava, Charles Gleason, PARIS Study Group, Juan Manuel Carreño, Viviana Simon, Florian Krammer

## Abstract

Mucosal immune responses are critical to prevent respiratory infections but it is unclear to what extent antigen specific mucosal secretory IgA (SIgA) antibodies are induced by mRNA vaccination in humans. We analyzed, therefore, paired serum and saliva samples from study participants with and without COVID-19 at multiple timepoints before and after severe acute respiratory syndrome coronavirus 2 (SARS-CoV-2) mRNA vaccination. Our results suggest that the level of mucosal SIgA responses induced by mRNA vaccination depend on pre-existing immunity. Indeed, vaccination induced only a weak mucosal SIgA response in individuals without pre-existing mucosal antibody responses to SARS-CoV-2 while SIgA induction after vaccination was efficient in COVID-19 survivors. Our data indicate that vaccinated seropositive individuals were able to swiftly induce relatively high anti-spike SIgA responses by boosting pre-existing mucosal immunity. In contrast, seronegative individuals did not have pre-existing anti-SARS-CoV-2 or cross-reacting anti-HCoV SIgA antibodies prior to vaccination, and, thus, little or no anti-SARS-CoV-2 SIgA antibodies were induced by vaccination in these individuals.

---

Severe acute respiratory syndrome coronavirus 2 (SARS-CoV-2), a novel betacoronavirus, was first identified in December 2019 as the causative agent of coronavirus disease 2019 (COVID-19). Since then, this virus has caused a pandemic that has so far claimed the life of more than 5.2 million people worldwide^1^. Since SARS-CoV-2 not only infects the epithelium of the lower respiratory tracts and causes pneumonia, but also the epithelium of upper respiratory tract^2^, secretory IgA (SIgA) antibodies may play an important role in the defense against infection. SIgA antibodies are IgA antibodies produced and transported from the mucosal stroma to the mucosal surface by utilizing the poly Ig receptor expressed on the basolateral side of epithelial cells^3^. Upon release of IgA antibodies to the mucosal surface, the extracellular segment of the poly Ig receptor, the secretory component (SC) is incorporated into the IgA molecules, which is a characteristic feature of SIgA antibodies. Another characteristic of SIgA antibodies is that these are mostly present in the form of multimers, such as dimers. These multimeric IgA antibodies display higher anti-viral activity than monomeric IgA antibodies^4^. SIgA antibodies are known to provide immediate immunity by eliminating respiratory pathogens before they pass through the mucosal barrier^5,6^. For example, during influenza virus infections, it has been reported that the main player in influenza immunity from previously infected individuals may be SIgA in the upper respiratory tract^7^. This adaptive immune response against influenza virus is primarily induced in secondary lymphoid tissues such as the mucosa-associated lymphoid tissue (MALT), which is responsible for the adaptive immune response induced at the virus infection and propagation site, and thus cannot be induced by injectable vaccines^3^. In the case of SARS-CoV-2, it has been reported that IgA antibodies that bind to the virus are rapidly produced, even before IgG antibodies, and can be detected in the serum and saliva of COVID-19 patients up to around 40 days post onset of symptoms as well^8–11^. However, it is unclear whether the currently used vaccines, including mRNA-based vaccines which are intramuscularly administered, induce SIgA antibody responses.

This study aims, thus, to determine whether antigen specific SIgA are induced in response to COVID-19 mRNA vaccination. We used longitudinal serum and saliva samples collected from 30 adult study participants over a period of 200 to 372 days. 18/30 of the study participants (60%) had COVID-19 and were seropositive prior to vaccination. 12/30 study participants (40%) had no previous SARS-CoV-2 infection history and were seronegative for SARS-CoV-2 antibodies prior to vaccination. Participants received either the Moderna mRNA-1273 vaccine or the Pfizer-BioNTech BNT162b2 vaccine. Demographics of study participants are summarized in **Supplementary Table 1**. Samples were collected at multiple timepoints prior to and after vaccination. We measured anti-SARS-CoV-2 spike binding IgG titers in serum samples and anti-SARS-CoV-2 spike IgG, SIgA and nucleoprotein (NP) SIgA titers in saliva by enzyme-linked immunosorbent assay (ELISA). One of the challenges in the measurement of antigen-specific SIgA titers in saliva samples is the fact that monomeric IgA and IgG antibodies from serum leak into saliva via crevicular fluid^3^. Therefore, a detection antibody that only recognizes human SC bound IgA antibodies was used to ensure specific detection of SIgA induced at the mucosa. Another challenge in the measurement of antigen-specific SIgA titers in saliva is that the IgA concentration within saliva is variable not only between individuals but within samples collected from the same individual, depending on various factors such as circadian rhythm, stress, and collection method of saliva samples^3^. We measured, therefore, the total IgA concentration within each saliva sample and normalized the SARS-CoV-2-specific SIgA titers based on the total IgA content within each saliva sample.

First, we evaluated the changes in anti-SARS-CoV-2 serum and saliva antibody titers before and after vaccination in each individual. An evident peak in anti-SARS-CoV-2 spike serum and mucosal antibody titers was observed in the seropositive group (**Fig.1** a, 1c, 1e). In the seronegative group, peaks in serum and saliva anti-spike IgG titers could be observed, though these titers were low compared to the seropositive group (**Fig.1** b and d). Interestingly, some individuals in the seronegative group presented a peak in anti-spike SIgA titers in saliva (**Fig.1** f). The peak that was observed in anti-SARS-CoV-2 spike reflected the mucosal IgA response to vaccination, as most individuals did not demonstrate a peak in anti-SARS-CoV-2 NP SIgA titer following vaccination (**Extended Data Fig.1**).

**Fig. 1.**
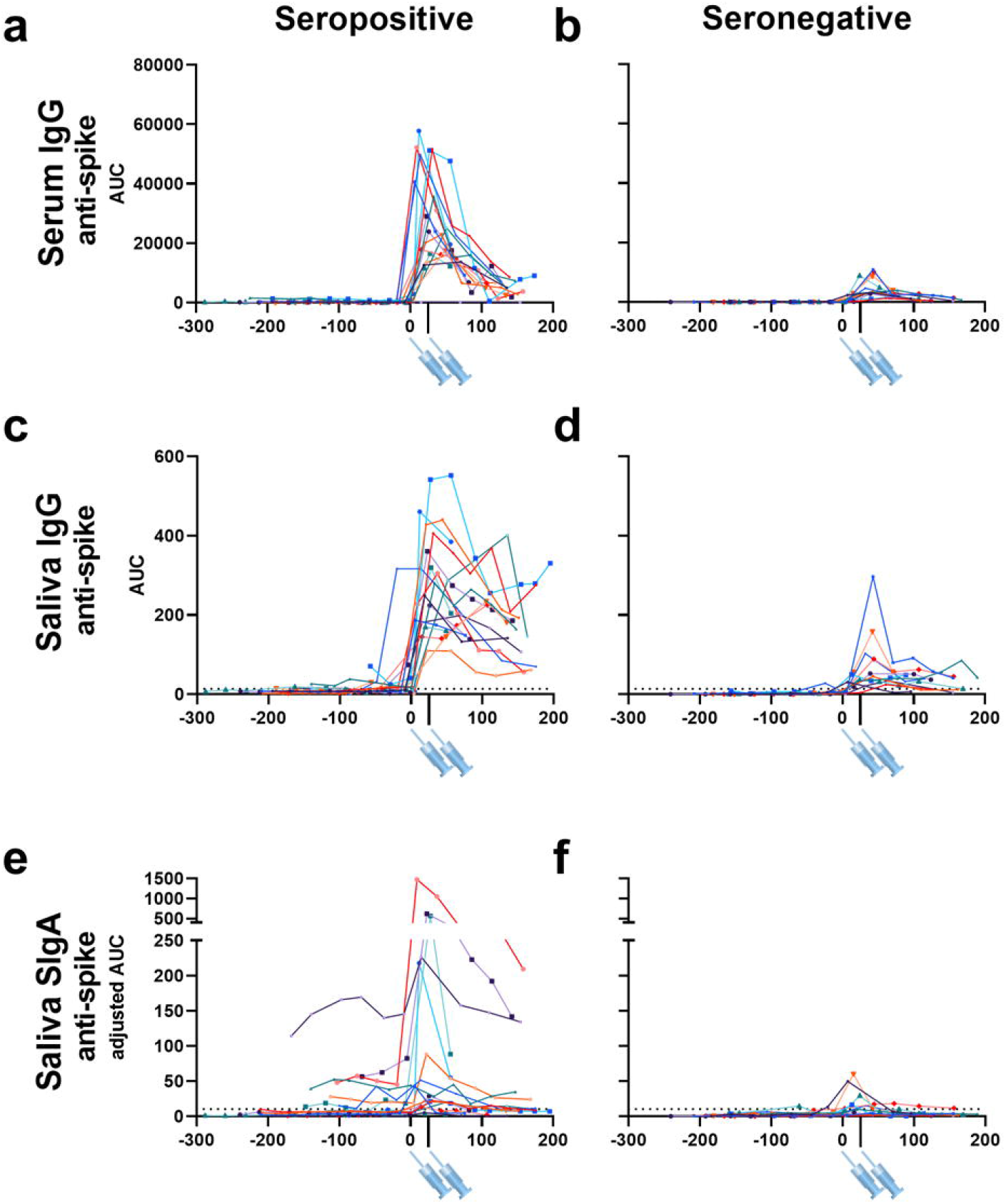
Time course of vaccine-induced serum and mucosal anti-SARS-CoV-2 antibody titers in adult participants with or without previous SARS-CoV-2 infection. Anti-SARS-CoV-2 spike IgG titers in serum (a, b), anti-SARS-CoV-2 spike IgG titers in saliva (c, d), and anti-SARS-CoV-2 spike SIgA titers in saliva (e, f) collected from 18 individuals with previous SARS-CoV-2 infection (seropositive) and 12 individuals without previous SARS-CoV-2 infection (seronegative) prior to and after vaccination. X-axis values represent days post first vaccination and Y-axis values represent antibody titers calculated as AUC (area under the curve). Syringe symbols point to the approximate time when the first and second vaccine dose were administered (28 days for Moderna mRNA-1273 vaccine and 21 days for the Pfizer-BioNTech BNT162b2 vaccine). Dotted lines are cut off values (mean+3SD of the AUC of samples from pre-vaccinated seronegative individuals). Different colors represent different individuals.

Next, we performed correlation analyses including all the samples tested to reveal the relationship between the different antibody titers. As expected, a positive correlation was observed between serum and saliva anti-spike IgG titers (**Fig. 2a**), reflecting the fact that the IgG antibodies measured in saliva were originally serum IgG antibodies that seeped into the mucosa from the circulation. In contrast, saliva anti-spike SIgA titers only weakly correlated with serum IgG titers (**Fig. 2b**). Next, to evaluate the difference in antibody titers over time, we grouped the data based on the collection timepoints: before vaccination, 1∼100 days post vaccination, and more than 100 days post vaccination. As anticipated serum IgG and mucosal SIgA antibody titers were significantly higher in seropositive individuals than seronegative individuals before vaccination (**Extended Data Fig.2a∼d**). The serum and mucosal antibody response to SARS-CoV-2 spike following vaccination remained significantly higher in seropositive individuals than seronegative individuals (**Extended Data Fig.2a∼c**). In contrast, the mucosal anti-NP SIgA antibody response was significantly higher in samples collected from previously infected individuals than those from previously naive individuals only before vaccination and 1∼100 days post vaccination. This difference was not observed in samples collected after 100 days post vaccination (**Extended Data Fig.2d**). This indicated that the anti-NP SIgA response observed in seropositive individuals was not boosted by vaccination, and weaned over time. Comparison between the different timepoints revealed that serum and mucosal anti-spike antibody titers significantly increased after vaccination in both seropositive and seronegative individuals (**Fig 2c and Extended Data Fig.2e∼f**). In contrast, increase in mucosal anti-NP IgA titers over time was not observed in either groups (**Extended Data Fig.2g**).

**Fig. 2.**
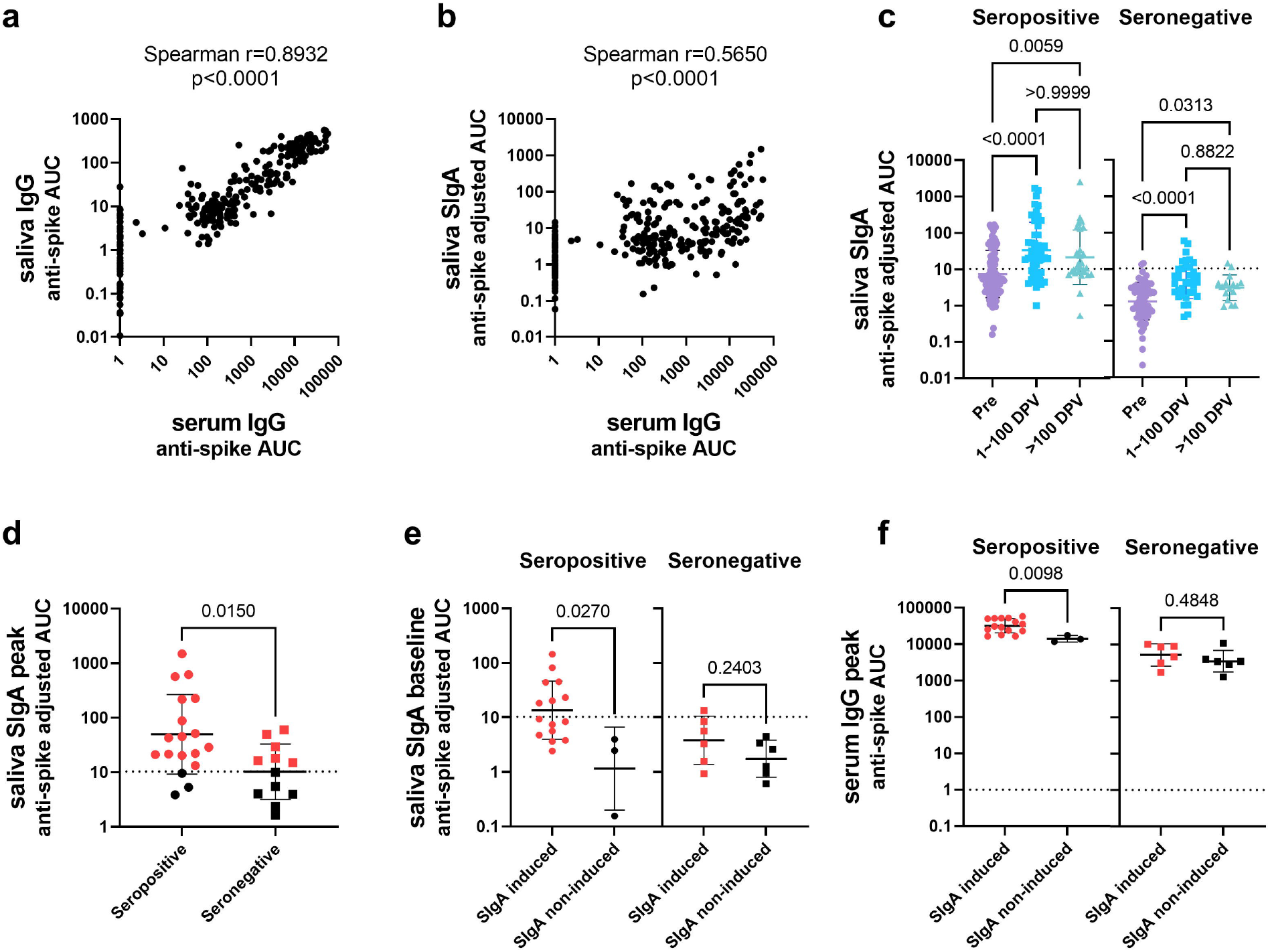
Mucosal SIgA antibody response to vaccination in adult participants with or without previous SARS-CoV-2 infection. (a, b) Correlation between anti-spike serum IgG antibody titers and saliva IgG antibody titers (a) or mucosal SIgA antibody titers (b) of samples collected from both previously infected and previously naïve individuals at all timepoints. X and Y-axis values represent antibody titers calculated as AUC. Spearman correlation coefficient and p values are labeled above each graph. (c) Comparison of anti-SARS-CoV-2 spike SIgA titers in saliva between samples collected during the three different timepoints: before vaccination (Pre), 1∼100 days post vaccination (1∼100 DPV), and over 100 days post vaccination (>100 DPV). Y-axis values are antibody titers calculated as AUC (area under the curve). Dotted lines are cut off values (mean+3SD of the AUC of samples from pre-vaccinated seronegative individuals). (One-way ANOVA with Kruskal-Wallis test). (d) Comparison of peak anti-SARS-CoV-2 spike mucosal SIgA titers between seropositive and seronegative individuals. The dotted line indicates the cut-off value (mean+3SD of the AUC of all samples collected from pre-vaccinated seronegative individuals). Red dots indicate SIgA induced individuals with peak SIgA titers above the cut-off value. (Mann-Whitney test) (e) Comparison of anti-SARS-CoV-2 spike SIgA titers in saliva of samples collected on the nearest possible date before vaccination (baseline) between individuals who presented mucosal anti-spike SIgA antibody titers above the cut-off value (SIgA induced) and those who did not (SIgA non-induced) in seropositive and seronegative individuals. The dotted line indicates the cut-off value (mean+3SD of the AUC of all samples collected from pre-vaccinated seronegative individuals). (Mann-Whitney test) (f) Comparison of peak anti-SARS-CoV-2 spike IgG titers in serum of SIgA induced or SIgA non-induced seropositive and seronegative individuals The dotted line indicates the cut-off value. (Mann-Whitney test).

In order to identify study participants who were able to successfully induce SIgA responses by vaccination, peak mucosal SIgA titers (the highest SIgA titer observed after vaccination) were assessed. Peak saliva SIgA titers were significantly higher in the seropositive individuals than the seronegative individuals (**Fig. 2d**). The same was observed for serum IgG titers (**Extended Data Fig.3a**), which was consistent with a previous report^12^. 15/18 seropositive individuals presented peak SIgA titers above the cut-off value (mean+3SD of the AUC for all samples collected from pre-vaccinated seronegative individuals), whereas only 6/12 seronegative individuals did (**Fig. 2d**). To identify factors associated with induction of SIgA by vaccination, baseline antibody titers (measured using samples collected on the nearest possible date before vaccination), were evaluated. In seropositive, anti-SARS-CoV-2 spike saliva SIgA titers were significantly higher in SIgA induced individuals (those with peak saliva SIgA titers above the cut-off value) than SIgA non-induced individuals (those with peak saliva SIgA titers below the cut-off value) (**Fig. 2e**). Significant differences were not observed in baseline anti-SARS-CoV-2 spike serum IgG and anti-SARS-CoV-2 NP saliva SIgA titers between SIgA induced and non-induced individuals (**Extended Data Fig.3b∼c**). However, for seronegative individuals, no significant difference was observed in baseline saliva anti-SARS-CoV-2 spike SIgA titers (**Fig. 2e**). It may be possible that mucosal immunity induced by seasonal coronaviruses (HCoVs) is boosted by vaccination and that this cross-reactive immunity (which mostly targets S2^13,14^, with cross-reactivity being common between betacoronaviruses OC43, HKU1 and SARS-CoV-2) is responsible for the weak induction of SIgA found in the previously seronegative individuals. To evaluate this, SIgA binding activity against the spike proteins of betacoronaviruses HCoV-OC43 and HCoV-HKU1, and the antigenically conserved SARS-CoV-2 S2 domain was assessed. However, no significant differences in baseline anti-HCoV-OC43, HCoV-HKU1, and SARS-CoV-2 S2 saliva SIgA titers between SIgA induced and non-induced groups was observed in seronegative individuals (**Extended Data Fig.3d**). In addition, the S2/RBD ratio of peak saliva SIgA was not significantly different between SIgA induced seropositive and seronegative individuals (**Extended Data Fig.3e**). However, the fold-increase of anti-SARS-CoV-2 spike SigA levels by vaccination was significantly higher than that of anti-HCoV-OC43 and HCoV-HKU1 spike SIgA titers in SIgA induced seropositive and seronegative individuals (**Extended Data Fig.3f**). These data indicate that the saliva SIgA antibodies induced by vaccination in seronegative individuals were not back-boosted antibodies against HCoVs, but rather SARS-CoV-2 spike specific antibodies that were newly elicited by vaccination. To assess whether the level of systemic immune response induced was associated with successful SIgA induction, peak serum anti-SARS-CoV-2 spike IgG antibodies were assessed. As a result, SIgA induced individuals presented significantly higher peak serum anti-SARS-CoV-2 spike IgG titers than SIgA non-induced individuals in seropositive individuals but not seronegative individuals. Interestingly, the peak serum anti-SARS-CoV-2 spike IgG titers observed in SIgA induced seronegative individuals was lower than SIgA non-induced seropositive individuals (**Fig. 2f**). Of note, neither the type of vaccine administered or sex did influence the induction of anti-SARS-CoV-2 spike saliva SIgA antibodies upon immunization (**Extended Data Fig.3g∼h**).

Our results support observations from other groups suggesting that intramuscular mRNA vaccination can induce SIgA antibodies in saliva^15,16^. Our results further suggest that vaccination alone can induce a weak mucosal SIgA response in individuals without pre-existing mucosal antibody response to SARS-CoV-2, although SIgA induction is much more efficient in individuals with pre-existing mucosal antibody response to SARS-CoV-2 elicited by COVID-19. Seropositive individuals presented significantly higher baseline anti-SARS-CoV-2 spike saliva SIgA titers in SIgA induced individuals than SIgA non-induced individuals, whereas baseline anti-SARS-CoV-2 spike saliva SIgA titers were not significantly different between SIgA induced and SIgA non-induced seronegative individuals. These observations suggested that vaccinated previously infected individuals were able to present a relatively quick and high anti-spike SIgA response by boosting the pre-existing mucosal anti-SARS-CoV-2 spike immunity. In contrast, seronegative individuals did not have pre-existing anti-SARS-CoV-2 or cross-reacting anti-HCoV SIgA antibodies prior to vaccination, and thus weak anti-SARS-CoV-2 SIgA antibodies were elicited by vaccination in 50% of the individuals. The mechanism by which intramuscularly administered vaccines induced a mucosal SIgA response remains unclear. Some hypotheses suggest that antigen presenting cells activated at the peripheral lymph nodes could migrate to the MALT and elicit a mucosal antibody response^17^. Alternatively, it has been reported that SARS-CoV-2 vaccine antigens have been detected in the plasma of the Moderna mRNA-1273 vaccinees^18^, suggesting that vaccine antigens may have reached the MALT to elicit mucosal immune responses. While we did not detect a correlation between seasonal betacoronavirus immunity and induction of SIgA in naïve vaccinated individuals, it could be that our methods were not sensitive enough and/or our sample size too small to discover a relationship. Thus, there is still a possibility that the SIgA responses observed here were a recall of immune memory elicited by past HCoV infections, which may cross-react with SARS-CoV-2. Peak serum anti-SARS-CoV-2 spike IgG titers were significantly higher in SIgA induced individuals than SIgA non-induced individuals in the previously infected group, indicating that the degree of immunological impact of vaccination on systemic immunity influences the boosting efficiency of pre-existing mucosal immunity. However, peak serum anti-SARS-CoV-2 spike IgG titers were not significantly different between SIgA induced and non-induced individuals in the previously non-infected group, and the peak serum anti-SARS-CoV-2 spike IgG titers observed in SIgA induced seronegative individuals was lower than SIgA non-induced seropositive individuals. This suggested that the degree of immunological impact of vaccination on systemic immunity does not have an influence on mucosal immunity priming but other factors determine the successful SIgA response in seronegative individuals.

Of note, among participants in the longitudinal observational PARIS (Protection Associated with Rapid Immunity to SARS-CoV-2) study, breakthrough infection cases after vaccination have so far only been identified in individuals without SARS-CoV-2 infection prior to vaccination, and not in individuals with SARS-CoV-2 infection prior to vaccination (personal communication, data not yet published). This, together with observations from the current study indicates that previously infected individuals mount mucosal SIgA antibody levels sufficient to provide protection from disease in response to vaccination. SIgA induction in individuals is expected to be beneficial in various aspects. First, it has been demonstrated that dimerization of IgA antibodies presented higher virus neutralization activity compared to monomeric IgA of anti-SARS-CoV-2 antibodies^19^. Second, IgA multimerization could increase cross-reactivity of antibodies, which may allow SIgA antibodies to provide protection against antigenic variant viruses^4,20^. Thus, vaccination strategies, such as intranasal vaccines like NDV-HXP-S, that could successfully induce SIgA should be sought for the control of the SARS-CoV-2 pandemic^21–23^. Further studies are needed to reveal the detailed mechanism of mucosal antibody induction by mRNA vaccination, to determine SIgA titers that would provide sterilizing immunity, and to evaluate the SIgA antiviral function in comparison to monomeric IgA.

## Data Availability

All data produced in the present study are available upon reasonable request to the authors.

## Acknowledgements

We thank the PARIS study participants for their generous and continued support. We would like to thank Kizzmekia Corbett and Barney Graham at the NIH VRC for sharing the HCoV spike expression plasmids. This work (cohort establishment, sample collection) is part of the PARIS/SPARTA studies funded by the NIAID Collaborative Influenza Vaccine Innovation Centers (CIVIC) contract 75N93019C00051.

This project has been funded in whole or in part with Federal funds from the National Cancer Institute, National Institutes of Health, under Contract No. 75N91019D00024, Task Order No.75N91021F00001. The content of this publication does not necessarily reflect the view s or policies of the Department of Health and Human Services, nor does mention of trade names, commercial products or organizations imply endorsement by the U.S. Government. K.S. is supported by the Japanese Society for the Promotion of Science (JSPS) Overseas Research Fellowship.

## Contributions

K.S. and F.K. conceived and designed the study. K.S., D.B., G.S., D.F., C.G., J.M.C., V.S., PVI study group and F.K. collected data, processed samples and selected samples. K.S. analyzed data. K.S. and F.K. wrote the manuscript and all the authors approved the final manuscript version.

## Extended data

**Extended Data Fig. 1.**
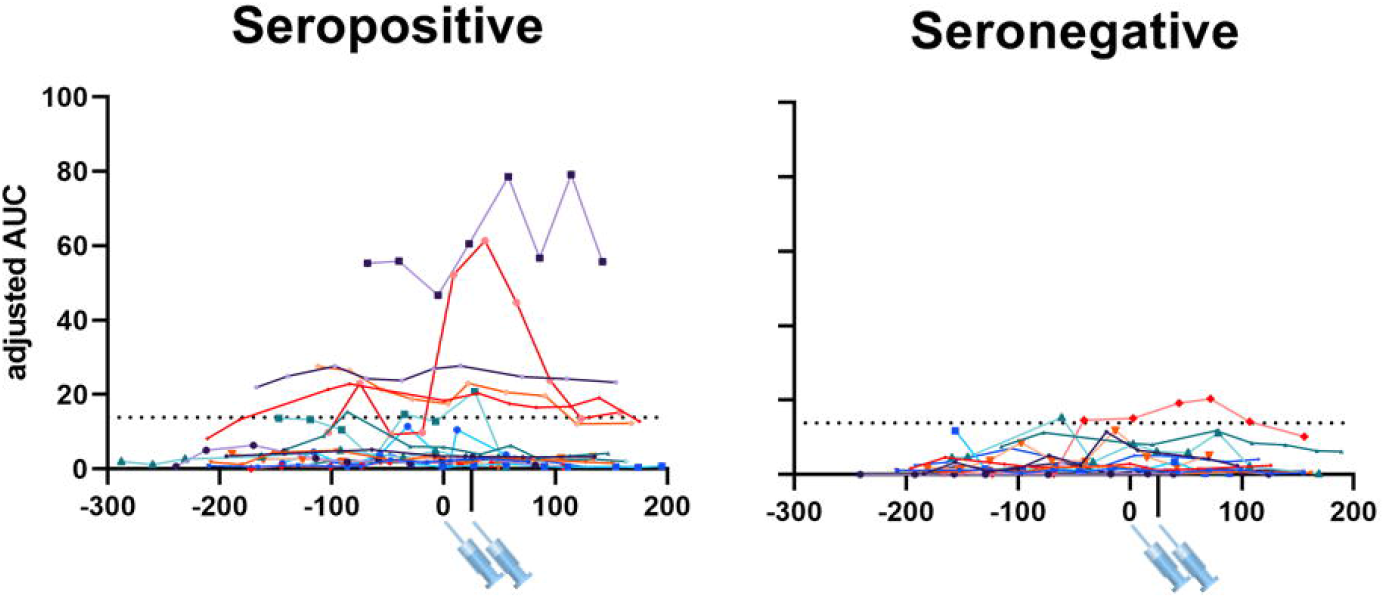
Mucosal antibody response to SARS-CoV-2 NP in humans with or without previous SARS-CoV-2 infection. Anti-SARS-CoV-2 NP SIgA titers in saliva collected from 18 individuals with previous SARS-CoV-2 infection (seropositive) and 12 individuals without previous SARS-CoV-2 infection (seronegative) prior to and after vaccination. X-axis values are days post first vaccination and Y-axis values are antibody titers calculated as AUC (area under the curve). Syringe symbols point to the approximate time when the first and second vaccine dose was administered (28 days for Moderna mRNA-1273 vaccine and 21 days for the Pfizer-BioNTech BNT162b2 vaccine). The dotted line indicates the cut-off value (mean+3SD of the AUC of samples from pre-vaccinated seronegative individuals). Different colors represent different individuals.

**Extended Data Fig. 2.**
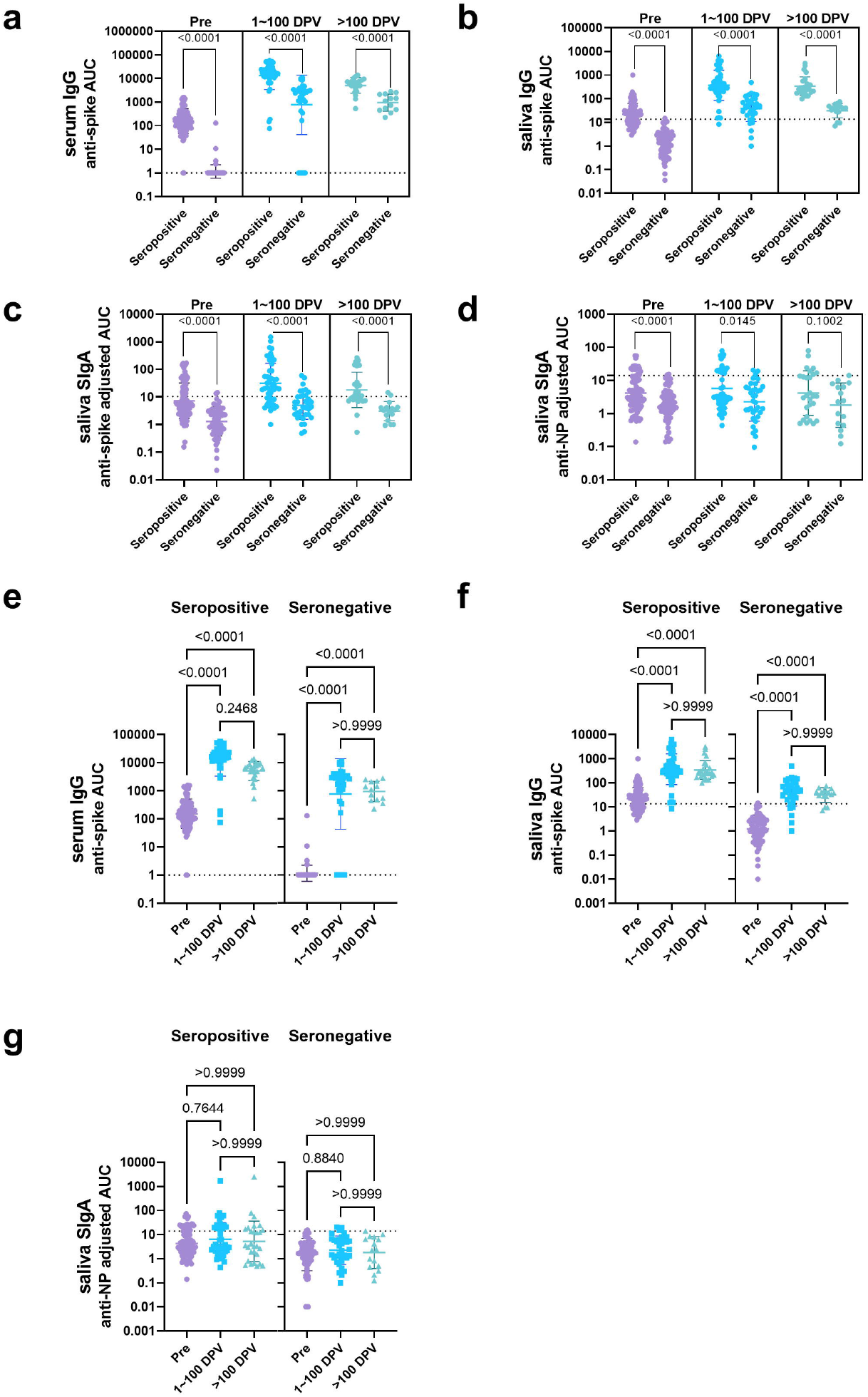
Comparison of serum and mucosal antibody titers between different timepoints. Comparison of anti-SARS-CoV-2 spike IgG titers in serum (a), anti-SARS-CoV-2 spike IgG titers in saliva (b), anti-SARS-CoV-2 spike SIgA titers in saliva (c), and anti-SARS-CoV-2 NP SIgA titers in saliva (d) between samples collected from seropositive and seronegative individuals. Samples were separated into three groups: those collected prior to vaccination (Pre, purple), at 1∼100 days post vaccination (1∼100 DPV, blue), and after 100 days post vaccination (>100 DPV, green) (Mann-Whitney test). Comparison of anti-SARS-CoV-2 spike IgG titers in serum (e), anti-SARS-CoV-2 spike IgG titers in saliva (f), and anti-SARS-CoV-2 NP SIgA titers in saliva (g) between samples collected during the three different timepoints. Dotted lines are cut off values (mean+3SD of the AUC of samples from pre-vaccinated seronegative individuals). (One-way ANOVA with Kruskal-Wallis test). Y-axis values are antibody titers calculated as AUC (area under the curve).

**Extended Data Fig. 3.**
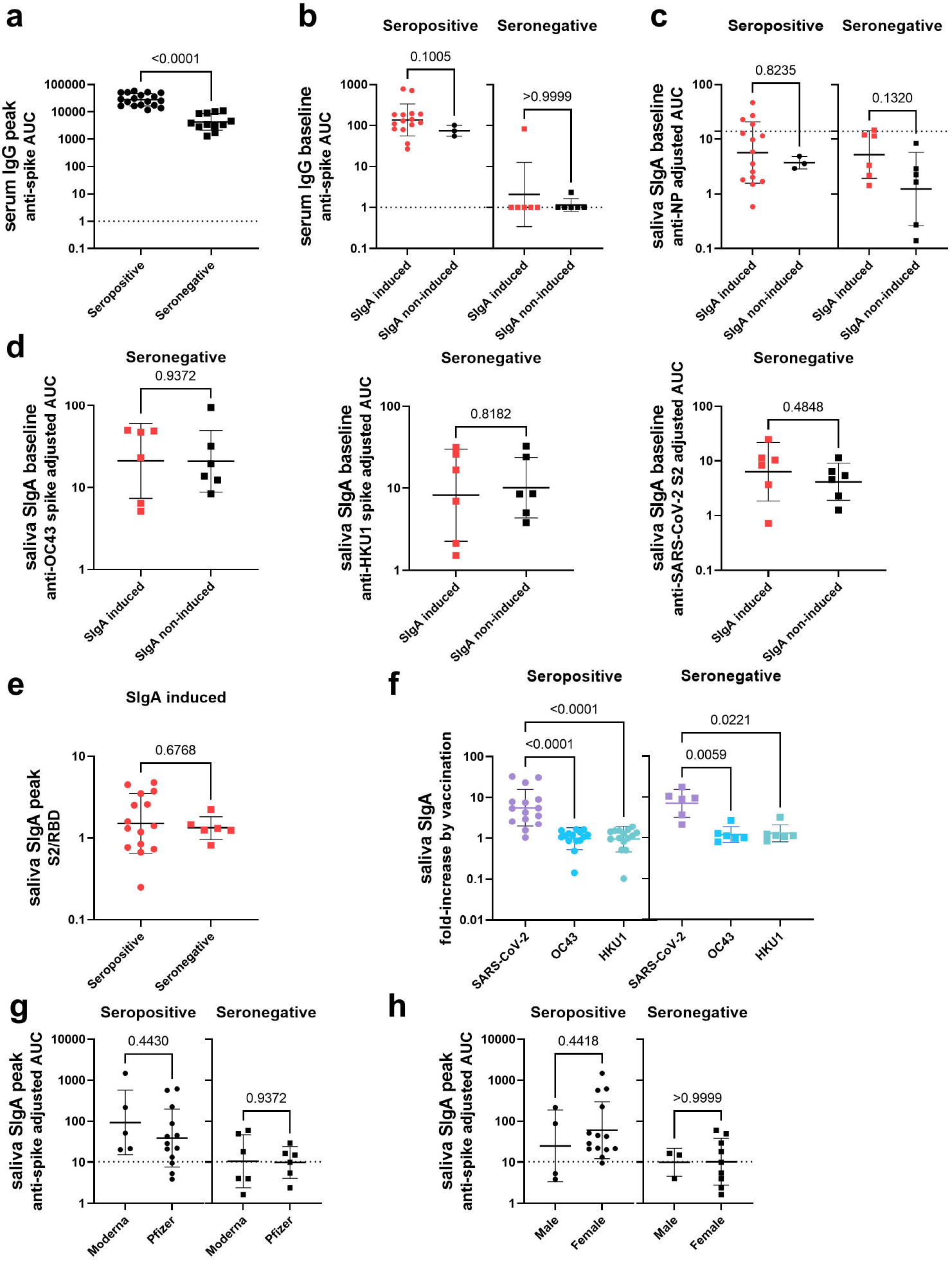
Analyses of antibody response to vaccination in the context of pre-existing antibody profiles and demographics of study participants. (a) Comparison of peak anti-SARS-CoV-2 spike serum IgG titers between seropositive individuals and seronegative individuals post vaccination (Mann-Whitney test). The dotted line indicates the cut-off value. (b, c) Comparison of anti-SARS-CoV-2 spike IgG titers in serum (b) and anti-NP SIgA titers in saliva (c) of samples collected on the nearest possible date before vaccination (baseline) between individuals who presented mucosal anti-spike SIgA antibody titers above the cut-off value (SIgA induced) and those who did not (SIgA non-induced) in seropositive and seronegative individuals (Mann-Whitney test). The dotted lines indicate the cut-off value (mean+3SD of the AUC of all samples collected from pre-vaccinated seronegative individuals for c). (d) Comparison of baseline anti-HCoV-OC43 spike (left panel), anti-HCoV-HKU1 spike (middle panel), and anti-SARS-CoV-2 spike S2 domain (right panel) reactivity between SIgA induced and SIgA non-induced seronegative individuals (Mann-Whitney test). (e) Comparison of peak saliva SIgA S2/RBD antibody ratio (calculated by dividing anti-SARS-CoV-2 S2 SIgA adjusted AUC by anti-SARS-CoV-2 RBD SIgA adjusted AUC) between seropositive and seronegative individuals who successfully induced SIgA in response to vaccination (Mann-Whitney test). (f) Comparison of fold increase in anti-SARS-CoV-2, anti-HCoV-OC43, and anti-HCoV-HKU1 SIgA titers in saliva by vaccination in seropositive and seronegative individuals. Fold increase was calculated by dividing peak SIgA adjusted AUC by baseline SIgA adjusted AUC. (One-way ANOVA with Kruskal-Wallis test) (g) Comparison of peak anti-SARS-CoV-2 spike SIgA titers between recipients of the Moderna mRNA-1273 vaccine (Moderna) and the Pfizer-BioNTech BNT162b2 vaccine (Pfizer) in seropositive and seronegative individuals (Mann-Whitney test). The dotted line indicates the cut-off value (mean+3SD of the AUC of all samples collected from pre-vaccinated seronegative individuals). (h) Comparison of peak anti-SARS-CoV-2 spike SIgA titers between male and female individuals (Mann-Whitney test). The dotted line indicates the cut-off value (mean+3SD of the AUC of all samples collected from pre-vaccinated seronegative individuals).

## Supplementary Information

**Supplementary Table 1:** Demographics of study participants and SIgA induction status

| Participant number | Age | Sex | Vaccine Type | Antibody status prior to vaccination | SIgA induction by vaccination |
| --- | --- | --- | --- | --- | --- |
| 1 | 36-40 | Male | Pfizer-BioNTech | positive | non-induced |
| 2 | 41-45 | Female | Moderna | negative | induced |
| 3 | 31-35 | Female | Pfizer-BioNTech | positive | induced |
| 4 | 56-60 | Male | Moderna | negative | non-induced |
| 5 | 31-35 | Female | Pfizer-BioNTech | positive | induced |
| 6 | 21-25 | Female | Moderna | positive | induced |
| 7 | 41-45 | Female | Pfizer-BioNTech | positive | induced |
| 8 | 26-30 | Female | Pfizer-BioNTech | negative | non-induced |
| 9 | 36-40 | Female | Pfizer-BioNTech | negative | non-induced |
| 10 | 31-35 | Female | Pfizer-BioNTech | positive | induced |
| 11 | 61-65 | Female | Moderna | negative | non-induced |
| 12 | 31-35 | Female | Pfizer-BioNTech | positive | induced |
| 13 | 36-40 | Female | Pfizer-BioNTech | negative | non-induced |
| 14 | 46-50 | Male | Pfizer-BioNTech | negative | induced |
| 15 | 41-45 | Male | Pfizer-BioNTech | positive | non-induced |
| 16 | 31-35 | Female | Pfizer-BioNTech | negative | induced |
| 17 | 36-40 | Female | Pfizer-BioNTech | positive | non-induced |
| 18 | 31-35 | Female | Pfizer-BioNTech | positive | induced |
| 19 | 31-35 | Female | Moderna | negative | induced |
| 20 | 26-30 | Female | Moderna | negative | induced |
| 21 | 36-40 | Female | Moderna | negative | non-induced |
| 22 | 21-25 | Female | Pfizer-BioNTech | positive | induced |
| 23 | 61-65 | Male | Pfizer-BioNTech | negative | induced |
| 24 | 31-35 | Female | Moderna | positive | induced |
| 25 | 46-50 | Female | Moderna | positive | induced |
| 26 | 36-40 | Male | Pfizer-BioNTech | positive | induced |
| 27 | 46-50 | Female | Moderna | positive | induced |
| 28 | 26-30 | Female | Pfizer-BioNTech | positive | induced |
| 29 | 46-50 | Male | Moderna | positive | induced |
| 30 | 41-45 | Female | Pfizer-BioNTech | positive | induced |

### Methods

#### Human samples (Participant enrollment, serum and saliva collection and methods)

Serum and saliva samples were collected from study participants in the longitudinal observational PARIS (Protection Associated with Rapid Immunity to SARS-CoV-2) study. The study was reviewed and approved by the Mount Sinai Hospital Institutional Review Board (IRB-20-03374). All participants signed written consent forms prior to sample and data collection. All participants provided permission for sample banking and sharing. Saliva was collected in a cup and serum by venipuncture. All biospecimen were coded and stored at -80C.

#### Recombinant proteins

Recombinant SARS-CoV-2 proteins were produced using a mammalian cell protein expression system. SARS-CoV-2 spike (S), receptor binding domain (RBD), and nucleoprotein (NP) gene sequences (GenBank: MN908947) were cloned into a mammalian expression vector, pCAGGs. The expression plasmids encoding for the spike of common human coronavirus (HCoV) OC43 and HKU1 were obtained from the NIH. S and RBD were produced as described previously^1,2^. Proteins were expressed using the Expi293 Expression System (Thermo Fisher Scientific), according to the manufacturer’s instructions. For S and RBD Proteins cell culture supernatants were collected and clarified by centrifugation at 4000 x g, filtered, and purified with Ni^2+^-nitrilotriacetic acid (NTA) Agarose (QIAGEN). For NP, cell pellets were collected, resuspended in high salt buffer (1M Tris-hydrochloric acid (HCl), pH 8.0; 0.5M ethylenediaminetetraacetic (EDTA) pH 8.0; 5M NaCl; Triton X-100 and distilled water) and lysed using the sonicator. Lysed cells were then centrifuged and supernatant was purified with Ni-NTA Agarose (QIAGEN). The purified proteins were concentrated using Amicon Ultracell (Merck) centrifugation units, and the buffer was changed to PBS (pH 7.4). Proteins were stored at -80°C until use. SARS-CoV-2 S2 protein was purchased from Sino Biological (#40590-V08B).

#### Antigen specific serum IgG ELISA

Serum IgG ELISA against SARS-CoV-2 spike was conducted as described previously^1,2^. Immulon 4 HBX 96-well microtiter plates (Thermo Fisher Scientific) were coated overnight at 4°C with recombinant proteins (100 ng/well) in PBS (pH 7.4). Well contents were discarded and blocked with 200 µL of 3% non-fat milk (AmericanBio) in PBS containing 0.1% Tween-20 (PBST) for one hour at room temperature (RT). After blocking, 100 µL of serum samples diluted (starting at 1:80 and serially diluted three-fold) with 1% non-fat milk in PBST was added to each well for reaction at RT for two hours. After washing with PBST three times, 50 μl of horseradish peroxidase (HRP)-labeled goat anti-human IgG antibody (Sigma-Aldrich, #A0170) diluted 3000-fold with 1% non-fat milk in PBST was added to each well, and incubated at RT for 1 hour. After washing with PBST three times, 100 μl of SIGMAFAST o-phenylenediamine dihydrochloride substrate solution (Sigma-Aldrich) was added to each well for the reaction at RT for 10 minutes. The reaction was stopped by addition of 50 μl of 3M HCl. Optical density at 490 nm was measured using Synergy 4 (BioTek) plate reader. Eight wells on each plate received no primary antibody (blank wells) and the optical density in those wells was used to assess background. Area under the curve (AUC) was calculated by deducting the average of blank values plus three times standard deviation of the blank values.

#### Antigen specific saliva SIgA and IgG ELISA

Immulon 4 HBX 96-well microtiter plates (Thermo Fisher Scientific) were coated overnight at 4°C with recombinant proteins (100 ng/well) in PBS (pH 7.4). Well contents were discarded and blocked with 200 µL of 5% non-fat milk (AmericanBio) in PBST for one hour at RT. After blocking, 50 µL of saliva samples diluted (starting at 1:2 and serially diluted two-fold) with 2.5% non-fat milk in PBST was added to each well. For IgG measurement, reaction of samples with antigen was conducted at RT for two hours. After washing with PBST three times, 50 μl of HRP-labeled goat anti-human IgG (H+L) antibody (Fisher Scientific, #PI31412) diluted to 0.32 mg/mL with 2.5% non-fat milk in PBST was added to each well, and incubated at RT for 1 hour. For SIgA measurement, reaction of samples with antigen was conducted at 4°C overnight. After washing with PBST three times, 50 μl of mouse anti-human secretory IgA antibody (MilliporeSigma, #HP6141) diluted to 5 μg/mL with 2.5% non-fat milk in PBST was added to each well, and incubated at RT for 2 hours. These plates were washed again with PBST three times, and 50 μl of HRP labeled goat anti-mouse IgG Fc antibody (Invitrogen, #31439) diluted to 1:1000 with 2.5% non-fat milk in PBST was added to each well, and incubated at RT for 1 hour. After washing with PBST three times, 100 μl of SIGMAFAST o-phenylenediamine dihydrochloride substrate solution (Sigma-Aldrich) was added to each well for the reaction at RT for 10 minutes. Reaction was stopped by addition of 50 μl of 3M HCl. Optical density at 490 nm was measured using Synergy 4 (BioTek) plate reader. Eight blank wells were used to assess background and AUC was calculated by deducting the average of blank values plus three times standard deviation of the blank values. Antigen specific SIgA AUC values were adjusted by dividing the values by total IgA concentration within saliva samples (adjusted AUC).

#### Measurement of total IgA concentration within saliva samples

Immulon 4 HBX 96-well microtiter plates (Thermo Fisher Scientific) were coated overnight at 4°C with 250 ng/well of goat anti-human IgA (Bethyl Laboratories, #A80-102A) in PBS (pH 7.4). Well contents were discarded and blocked with 200 µL of 5% non-fat milk (AmericanBio) in PBST for one hour at RT. After blocking, 50 µL of serum samples diluted (starting at 1:256 and serially diluted three-fold) with 2.5% non-fat milk in PBST was added to each well and incubated at RT for two hours. After washing with PBST three times, 50 μl of HRP-labeled goat anti-human IgA antibody (Bethyl Laboratories, #A80-102P) diluted to 1:10000 with 2.5% non-fat milk in PBST was added to each well, and incubated at RT for 1 hour. After washing with PBST three times, 100 μl of SIGMAFAST o-phenylenediamine dihydrochloride substrate solution (Sigma-Aldrich) was added to each well for the reaction at RT for 10 minutes. Reaction was stopped by addition of 50 μl of 3M hydrochloric acid (HCl). Optical density at 490 nm was measured using Synergy 4 (BioTek) plate reader. Purified human plasma IgA (EMD Millipore, #401098) was diluted (starting at 500 ng/mL and serially diluted two-fold) was used as standard. A five-parameter logistic fit was conducted on the standard curve and IgA concentrations within saliva samples were calculated.

#### Statistical analysis

Differences between antibody titers between two groups was analyzed with Mann-Whitney test. Differences between antibody titers between three groups was analyzed with one-way ANOVA with Kruskal-Wallis. Correlations between antibody titers were analyzed using Spearman’s rank test. All statistical analyses, AUC calculation, and five-parameter logistic fit of standard curves were conducted with Graphpad Prism version 9.

